# Evaluation of 30-day mortality for 500 patients undergoing non-emergency surgery in a COVID-19 cold site within a multicentre regional surgical network during the COVID-19 pandemic

**DOI:** 10.1101/2020.06.10.20115543

**Authors:** Veeru Kasivisvanathan, Jamie Lindsay, Sara Rakshani-Moghadam, Ahmed Elhamshary, Konstantinos Kapriniotis, Georgios Kazantzis, Bilal Syed, John Hines, Axel Bex, Daniel Heffernan Ho, Martin Hayward, Chetan Bhan, Nicola MacDonald, Simon Clarke, David Walker, Geoff Bellingan, James Moore, Jennifer Rohn, Asif Muneer, Lois Roberts, Fares Haddad, John D Kelly, UCLH study group collaborators

## Abstract

**Background:** Two million non-emergency surgeries are being cancelled globally every week due to the COVID-19 pandemic, which will have a major impact on patients and healthcare systems.

**Objective:** To determine whether it is feasible and safe to continue non-emergency surgery in the COVID-19 pandemic

**Design, setting and participants:** This is a cohort study of 500 consecutive patients undergoing non-emergency surgery in a dedicated COVID-19 cold site following the first case of COVID-19 that was reported in the institution. The study was carried out during the peak of the pandemic in the United Kingdom, which currently has one of the highest number of cases and deaths from COVID-19 globally.

We set up a hub-and-spoke surgical network amongst 14 National Health Service institutions during the pandemic. The hub was a cancer centre, which was converted into a COVID-19 cold site, performing urological, thoracic, gynaecological and general surgical operations.

**Outcomes:** The primary outcome was 30-day mortality from COVID-19. Secondary outcomes included all-cause mortality and post-operative complications at 30-days.

**Results:** 500 patients underwent surgery with median age 62.5 (IQR 51-71). 65% were male and 60% had a known diagnosis of cancer. 44% of surgeries were performed with robotic or laparoscopic assistance and 61% were considered complex or major operations.

None of the 500 patients undergoing surgery died from COVID-19 at 30-days. 30-day allcause mortality was 3/500 (1%). 10 (2%) patients were diagnosed with COVID-19, 4 (1%) with confirmed laboratory diagnosis and 6 (1%) with probable COVID-19. 33/500 (7%) of patients developed Clavien-Dindo grade 3 or higher complications, with 1/33 (3%) occurring in a patient with COVID-19.

**Conclusion:** It is safe to continue non-emergency surgery during the COVID-19 pandemic with appropriate service reconfiguration.

**Patient summary:** No patients died from COVID-19 when undergoing non-emergency surgery during the pandemic in one of the worst affected world regions.

## Background

COVID-19 has led to most non-emergency surgery in regions affected by the COVID-19 pandemic being halted [1] in an effort to divert resources and staff to managing patients with COVID-19 and to reduce the impact of COVID-19 on patients undergoing surgery. Globally, it is estimated that over 2 million non-emergency operations are being cancelled each week due to COVID-19 [2], This will have a profoundly detrimental long-term effect on patients and healthcare systems. Patients’ quality of life and survival can be reduced by delayed surgery and there are significant health economic consequences to the population [3-6].

An international cohort study reported a concerning 19% 30-day mortality in 278 patients undergoing non-emergency surgery who had COVID-19 diagnosed peri-operatively[7]. There are a number of mechanisms by which surgery may result in worse outcomes for those infected with COVID-19. Surgery is known to impair immune function [8], can lead to a dysregulated inflammatory response [9] and can lead to a high incidence of respiratory complications [7,10].

The UK is globally one of the worst-affected countries from COVID-19, with over 259,559 confirmed cases and 36,793 deaths as of the 24^th^ May 2020[11]. The first case in the UK was recorded on the 30^th^ January 2020 and London is the UK region with the highest number of reported cases [11]. In order to continue to safely provide a surgical service to patients who would benefit from their urgent cancer surgery, we set up a multicentre surgical network based in the London area, taking regional and national referrals for urgent surgery and performing these surgeries centrally at a site that was intended to be kept a COVID-free site during the COVID-19 pandemic. This was part of an approach coordinated by the Pan-London Cancer Alliances and NHS England.

We aimed to assess the 30-day mortality rate from COVID-19 in patients undergoing nonemergency surgery at our institution during the peak of the pandemic. We hoped to demonstrate that it can be both feasible and safe to continue with the conduct of nonemergency surgery.

## Methods

### Study design

This was a cohort study evaluating patients undergoing non-emergency surgery at a dedicated COVID-19 cold site (CCS), within a regional urgent surgery network of 14 National Health Service hospital trusts. The study is reported according to the Strengthening the Reporting of Observational Studies in Epidemiology guidelines [12].

### Setting

Our institution consists of a number of geographically separate sites located within a 2-mile distance in London, the region with the highest number of confirmed cases in the UK [11], We converted one of these sites, which was a high volume urological and thoracic cancer centre, into a dedicated CCS. This CCS has 7 operating theatres, 84 inpatient beds and a level-1 surgical ITU with 9 beds. The aim of the service restructuring within our institutional sites was to maximise the chances of keeping the dedicated CCS COVID-19 free and keep urgent cross-speciality surgery going (Table 1) [13].

**Table 1:** Healthcare service restructuring in response to COVID-19

| Type of restructuring | Description |
| --- | --- |
| Regional referral network | • Organisation of cancer and urgent surgery network consisting of 14 UK National Health Service Trusts (University College London Hospital NHS Foundation Trust, Royal Free London NHS Foundation Trust, North Middlesex University Hospital NHS Trust, Barts Health NHS Trust, Whittington Health NHS Trust, Cambridge University Hospitals NHS Foundation Trust, United Lincolnshire Hospitals NHS Trust, University Hospital Southampton NHS Foundation Trust, Barking, Havering and Redbridge University Hospitals NHS Trust, The Princess Alexandra Hospital NHS Trust, Homerton University Hospital NHS Foundation Trust, Bedfordshire Hospitals NHS Foundation Trust, Ashford and St Peter's Hospital NHS Foundation Trust and the Royal Brompton and Harefield NHS Foundation Trust)<br>• Network arranged into Hub-and-Spoke organisational design[13] where the anchor site and hub for conducting the major urological, thoracic, gynaecological and general surgery was a dedicated COVID-19 cold site<br>• Patients with an urgent need for surgery from the remaining regional and national network sites (spokes) were referred for surgery at the dedicated COVID-19 cold site hub.<br>• Surgeons from local referring institutions were set up with operating rights at the cold site hub and could perform surgery on the patients they had referred. |
| Reconfiguration across institutional sites | • Creation of COVID-19 hot and cold sites within our institution. Unwell patients with suspected COVID-19 were admitted only to hot sites. Conversion of one of our institutional sites into a dedicated COVID-19 cold site. Non-emergency surgery that would typically occur prior to the COVID-19 pandemic in the hot sites were diverted to the cold site during the pandemic.<br>• No emergency admissions or direct patient transfers were accepted at the COVID-19 cold site during the COVID-19 period for urological, gynaecological or general surgery. It was mandated that any transfers or emergencies in in these specialties were admitted directly to the hot sites.<br>• Though the clinicians managing these patients at hot sites were based in the cold site, a dedicated sub-team attended the hot site evaluating and managing and the patients admitted there.<br>• In thoracic surgery due to the urgent nature of the pathology, urgent transfers were accepted to the cold site, but only if they had a negative COVID-19 viral swab prior to transfer. |
| Reconfiguration at hub COVID-19 cold site where surgery was performed | • Staff were set up with remote access to the electronic record system<br>• Outpatient services were converted from face-to-face appointments to telephone appointments where feasible |
|  | • Administrative and clinical staff worked from home where feasible<br>• Multi-disciplinary team meetings were carried out by web conferences, with a restriction placed on the maximum numbers of attendees for essential face-to-face meetings to 5 people<br>• Staff treating inpatients on the wards were required to wear a surgical mask, an apron and a pair of gloves for each patient<br>• Family members were not allowed to visit inpatients<br>• Patients were called before surgery to ensure they were asymptomatic<br>• Patients were asked to self-isolate, where feasible, 14 days prior to and after their surgery |
| Reconfiguration of the theatre environment | • Full personal protective equipment worn by each member of staff included an apron, surgical gown, two pairs of gloves, F95 mask, face visor and theatre hat.<br>• Dedicated areas for donning and doffing were created, training was provided on performing these manoeuvres, and a dedicated donning team assisted each member of staff.<br>• The patient would be intubated and extubated in theatre with only the anaesthetist and operating department practitioner present. After intubation and extubation, other staff did not enter the theatre for 20 minutes to minimise risk of exposure to aerosolised airway secretions.<br>• During surgery the number of staff in theatre was kept to the minimum required.<br>• The number of planned cases on each theatre list was reduced in order to facilitate longer turnaround time between cases.<br>• During robotic surgery, a smoke evacuation device was used for all cases to minimise the putative risk of transmission of COVID-19 virus particles into the theatre environment. |

### Patients

The first 500 consecutive patients having non-emergency surgery at the CCS from the 5^th^ March 2020 (the date of first case of COVID-19 in our institution) to 22^nd^ April 2020 were included. On the 26^th^ March 2020 a regional cancer and urgent surgery network was set up with representation from urology, thoracic, gynaecology and general surgery (Figure 1). This allowed patients from other institutions and other specialities in the network with the greatest need for urgent surgery to have this at the CCS. In urological surgery, non-urgent and non-cancer surgery stopped after inception of the regional network. Patients were prioritised, influenced by national guidelines, on basis of their individual cancer risk and potential benefit of having surgery [3,14] judged against patient risk for serious complications of COVID-19 [15]. In thoracic surgery, due to the urgent nature of the surgery, elective cancer and urgent surgery continued unabated.

**Figure 1.**
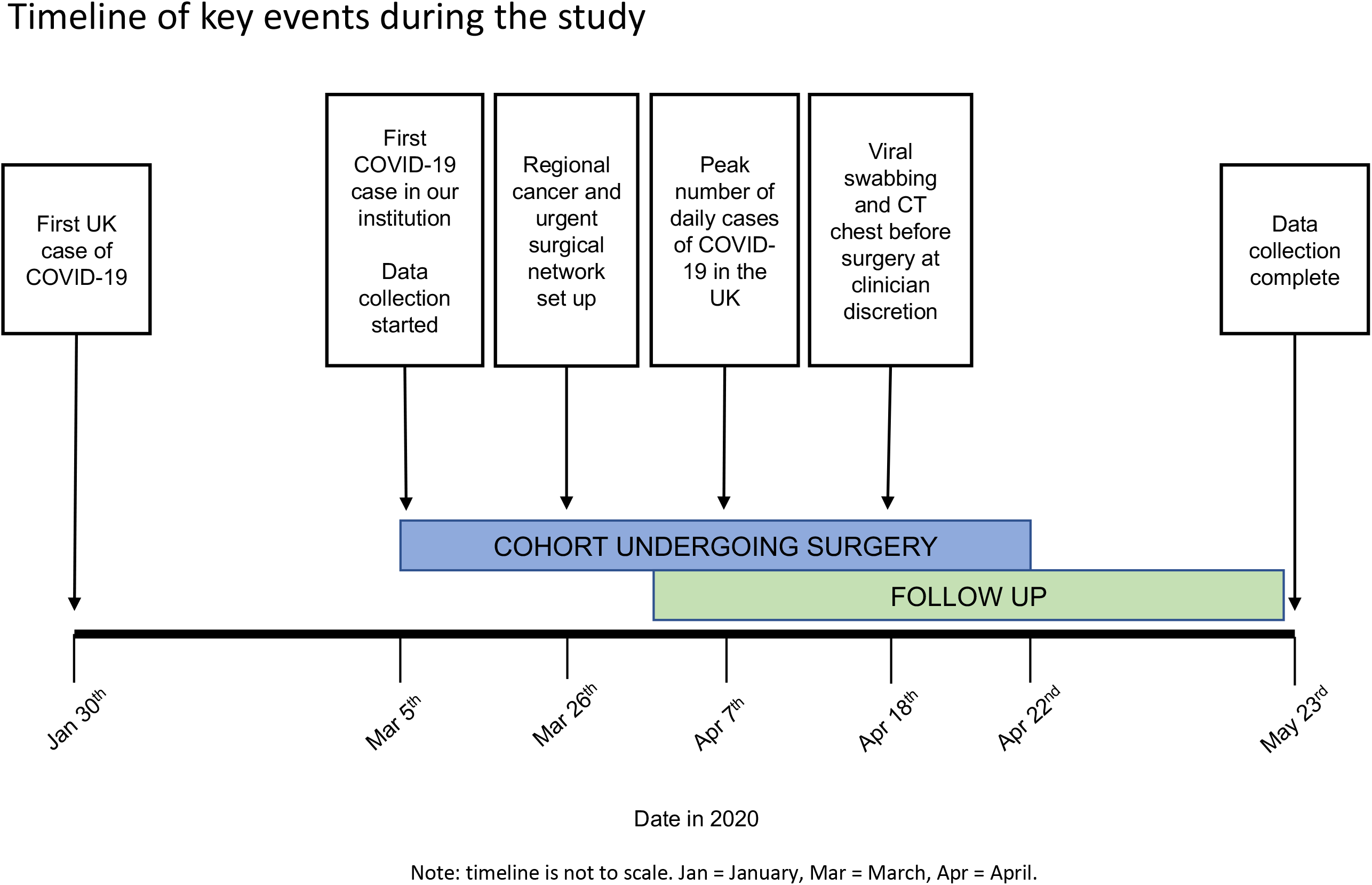
Timeline of key events during the study. Note: timeline is not to scale. Jan = January, Mar = March, Apr = April.

### Primary outcome

The primary outcome was the proportion of patients who died from COVID-19 within 30-days of surgery. Cause of death was assessed by the clinical care team and were extracted from death certificates, following national guidelines [16].

### Secondary outcomes

Secondary outcomes included the proportion of patients who died from any cause within 30-days, the proportion of patients developing confirmed or probable COVID-19 within 30-days and the 30-day post-operative complication profile.

The date of onset of COVID-19 was defined as the date on which the first related symptoms appeared. In patients undergoing testing, the presence of COVID-19 RNA was assessed with a real-time reverse transcriptase polymerase chain reaction technique on a nasopharyngeal and oropharyngeal swab collected according to World Health Organisation (WHO) recommendations [17], utilizing the Hologic Panther Fusion assay.

In line with WHO guidelines, a diagnosis of confirmed COVID-19 was given to patients with laboratory confirmation of COVID-19 infection, irrespective of clinical signs and symptoms[18]. A diagnosis of probable COVID-19 was given to patients who did not undergo laboratory testing or whose laboratory testing was inconclusive, but who had fever and at least one sign of acute respiratory illness (persistent cough, shortness of breath, sore throat, loss of smell, loss of taste or vomiting). The proportion of patients with a chest CT with the typical appearances of COVID-19 pneumonia according to the Radiological Society of North America was also reported [19]. Surgical complications were graded according to the Clavien-Dindo classification [20].

### Surgical precautions

Patients were called prior to the day of their operation and were only asked to attend for surgery if they remained asymptomatic. Where feasible, patients were asked to self-isolate for 14 days prior to their surgery.

From 6^th^ April 2020, in line with national recommendations, staff wore personal protective equipment (PPE) and took precautions assuming as default that the patient had unrecognised COVID-19 infection[21] (Table 1).

**Table 2:**
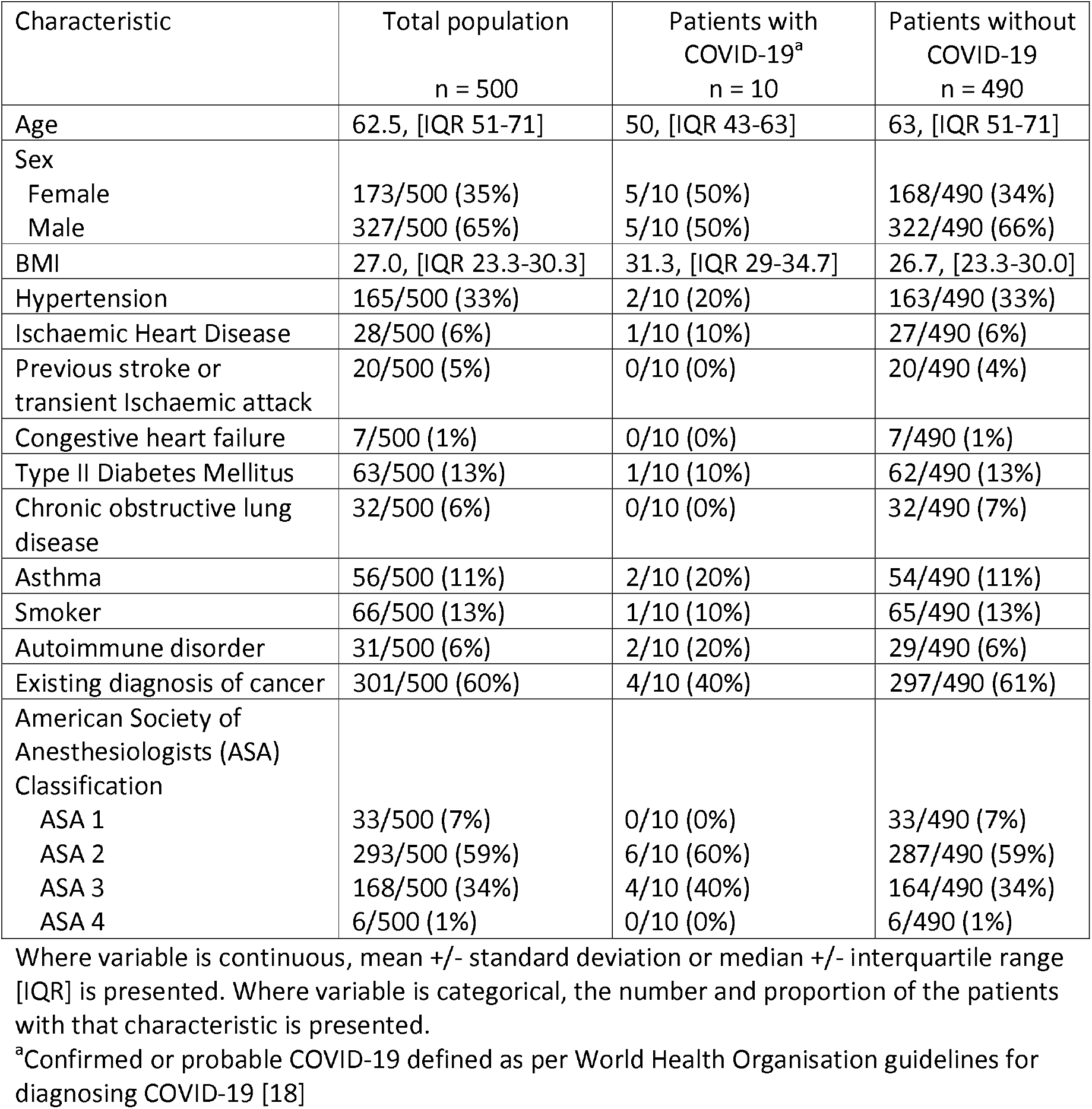
Baseline demographics of all patients undergoing surgery, patients diagnosed with COVID-19 and patients who did not develop COVID-19

| Characteristic | Total population<br>n = 500 | Patients with<br>COVID-19<sup>a</sup><br>n = 10 | Patients without<br>COVID-19<br>n = 490 |
| --- | --- | --- | --- |
| Age | 62.5, [IQR 51-71] | 50, [IQR 43-63] | 63, [IQR 51-71] |
| Sex |  |  |  |
| Female | 173/500 (35%) | 5/10 (50%) | 168/490 (34%) |
| Male | 327/500 (65%) | 5/10 (50%) | 322/490 (66%) |
| BMI | 27.0, [IQR 23.3-30.3] | 31.3, [IQR 29-34.7] | 26.7, [23.3-30.0] |
| Hypertension | 165/500 (33%) | 2/10 (20%) | 163/490 (33%) |
| Ischaemic Heart Disease | 28/500 (6%) | 1/10 (10%) | 27/490 (6%) |
| Previous stroke or<br>transient Ischaemic attack | 20/500 (5%) | 0/10 (0%) | 20/490 (4%) |
| Congestive heart failure | 7/500 (1%) | 0/10 (0%) | 7/490 (1%) |
| Type II Diabetes Mellitus | 63/500 (13%) | 1/10 (10%) | 62/490 (13%) |
| Chronic obstructive lung<br>disease | 32/500 (6%) | 0/10 (0%) | 32/490 (7%) |
| Asthma | 56/500 (11%) | 2/10 (20%) | 54/490 (11%) |
| Smoker | 66/500 (13%) | 1/10 (10%) | 65/490 (13%) |
| Autoimmune disorder | 31/500 (6%) | 2/10 (20%) | 29/490 (6%) |
| Existing diagnosis of cancer | 301/500 (60%) | 4/10 (40%) | 297/490 (61%) |
| American Society of<br>Anesthesiologists (ASA)<br>Classification |  |  |  |
| ASA 1 | 33/500 (7%) | 0/10 (0%) | 33/490 (7%) |
| ASA 2 | 293/500 (59%) | 6/10 (60%) | 287/490 (59%) |
| ASA 3 | 168/500 (34%) | 4/10 (40%) | 164/490 (34%) |
| ASA 4 | 6/500 (1%) | 0/10 (0%) | 6/490 (1%) |

Where variable is continuous, mean +/- standard deviation or median +/- interquartile range [IQR] is presented. Where variable is categorical, the number and proportion of the patients with that characteristic is presented.
<sup>a</sup>Confirmed or probable COVID-19 defined as per World Health Organisation guidelines for diagnosing COVID-19 [18]

From 18^th^ April 2020, at the discretion of their treating clinician, patients underwent COVID-19 viral swab testing and CT of the chest 48 hours before their surgery if they were planned for ITU admission post-operatively or were deemed by their clinical team to be high risk for complications of COVID-19.

### Post-operative management

Patients were evaluated on daily ward rounds during their inpatient stay. If patients presented with symptoms consistent with COVID-19 they were isolated in a side room and tested for COVID-19 with a viral swab and chest CT.

Once discharged, patients were instructed to self-isolate for 14 days where feasible. A phone call at or shortly after 30 days was carried out to determine their clinical status.

### Data collection

We reviewed electronic medical records with a standardised case report form. We assessed baseline demographics, operation notes, radiological test results, laboratory test results and post-operative clinical encounters. Data entry was verified independently by two individuals.

### Statistical analysis

Continuous data were presented with mean and standard deviation or median and interquartile range. Categorical data were presented with the number of patients and percentage in each category. All analyses were performed using STATA (version 14.2) software.

### Ethics

The institutional review board at University College London Hospital deemed this work exempt from ethical approval.

## Results

The first confirmed case of COVID-19 in our institution was on 5^th^ March 2020. 500 patients underwent non-emergency surgery at the dedicated CCS between the 5^th^ March 2020 and 22^nd^ April 2020. The date of follow up for the final patient was on the 23^rd^ May 2020. In this time period, across all of our institutional sites, there were 788 confirmed cases of COVID-19.

The median hospital inpatient stay was 1 night. Patients were of median age 62.5 and 65% were male (Table 2). 350/500 (70%) of operations were performed for the diagnosis or treatment of cancer and 150/500 (30%) were done for urgent non-cancer or benign conditions (Table 3).

**Table 3:** A table showing the surgeries performed classified by speciality, complexity and number performed

| Speciality and operation, stratified by complexity of surgery<sup>a</sup> | Number performed (%)<br>N = 500 |
| --- | --- |
| **Urology** | **N = 333/500 (67%)** |
| *Major or complex* | *n = 160* |
| Excision of penile/perineal lesion and graft | 3 |
| Glansectomy +/- graft for penile cancer | 3 |
| Insertion of artificial urethral sphincter | 5 |
| Insertion or removal of penile prosthesis | 3 |
| Radical nephrectomy or nephroureterectomy | 13 |
| Radical cystectomy and/or urinary diversion | 19 |
| Radical prostatectomy | 45 |
| Radical penectomy | 3 |
| Urethroplasty | 5 |
| Transurethral resection of bladder tumour | 13 |
| Ureterorenoscopy +/- procedure | 26 |
| Other major surgery | 22 |
| *Intermediate* | *n = 95* |
| Cryotherapy to prostate | 10 |
| High intensity focal ultrasound of the prostate | 10 |
| Insertion or exchange of nephrostomy | 10 |
| Radical orchidectomy | 1 |
| Rigid cystoscopy +/- procedure | 56 |
| Other intermediate surgery | 8 |
| *Minor* | *n = 78* |
| Circumcision for penile cancer | 11 |
| Flexible cystoscopy +/- procedure | 18 |
| Insertion of suprapubic catheter | 5 |
| Penile biopsy | 1 |
| Transperineal prostate biopsy | 31 |
| Other minor surgery | 12 |
| **Thoracics** | **N = 117/500 (23%)** |
| *Major or complex* | *n = 107* |
| Lobectomy | 26 |
| Excision of lung lesion | 38 |
| Video assisted thoracoscopic procedure | 39 |
| Other major surgery | 4 |
| *Intermediate* | n = 10 |
| Bronchoscopy | 3 |
| Mediastinoscopy | 4 |
| Insertion of chest drain | 3 |
| Gynaecology | N = 45/500 (9%) |
| *Major or complex* | n = 34 |
| Total abdominal hysterectomy +/- bilateral salpingoopherectomy | 31 |
| Other major surgery | 3 |
| *Intermediate* | n = 5 |
| Evacuation of retained products of conception | 4 |
| Loop excision of transformation zone | 1 |
| *Minor* | n = 6 |
| Hysteroscopy | 2 |
| Other minor surgery | 4 |
| General surgery | N = 5/500 (1%) |
| *Major* | n = 4 |
| Adrenalectomy | 1 |
| Bowel resection | 1 |
| Haemorrhoidectomy | 1 |
| Thyroidectomy | 1 |
| *Minor* | n = 1 |
| Examination of rectum under anaesthesia | 1 |

<sup>a</sup>Complexity as per NICE guidelines [NG45]: Routine preoperative tests for elective surgery[22]

220/500 (44%) of operations were performed with robotic or endoscopic assistance, with the remaining performed via an open, percutaneous or natural orifice approach. 305/500 (61%) were classified as major or complex surgery, 110/500 (22%) as intermediate and 85/500 (17%) as minor[22]. Pre-operatively, 72/500 (14%) patients underwent preoperative viral swabs and 22/500 (4%) underwent pre-operative chest CT. Of these none had a laboratory confirmed test result positive for COVID-19 though one patient had changes with typical appearances of COVID-19 on chest CT. This patient was asymptomatic and had probable COVID-19 infection one month prior. In light of the CT changes, this patient’s surgery was deferred by two weeks but was performed during the study.

No patient died from COVID-19 at 30-days. The all cause 30-day mortality was 3/500 (1%). Causes of death included aspiration pneumonia secondary to small bowel obstruction, myocardial infarction in a patient with underlying ischaemic heart disease and metastatic breast cancer. The latter two deaths occurred after the patients had been discharged home. 10/500 (2%) patients were diagnosed with confirmed or probable COVID-19 (Table 2), of whom 4/500 (1%) were confirmed on a viral swab (Table 4). 6/500 (1%) patients were diagnosed with probable COVID-19, with fever and at least one sign of acute respiratory illness.

**Table 4:** The diagnosis of COVID-19 in 500 patients undergoing surgery at a dedicated COVID-19 cold site

| Characteristic | Summary measure |
| --- | --- |
| *Pre-operative* |  |
| Number of patients with pre-operative viral swab sent off for COVID-19 | 72/500 (14%) |
| Number of patients with a pre-operative viral swab positive for COVID-19 | 0/72 (0%) |
| Number of patients with pre-operative CT chest | 22/500 (3%) |
| Number of patients with pre-operative CT chest with changes typical of COVID-19<sup>a</sup> | 1/22 (5%) |
| *Post-operative* |  |
| Number of patients with post-operative viral swabs sent off for COVID-19 | 41/500 (8%) |
| Number of viral swabs sent off post-operatively for COVID-19 | 44 |
| Median number of days from surgery to post-operative viral swab for COVID-19 (median, IQR) | 5 [IQR 2-12] |
| Number of patients undergoing post-operative chest CT | 19/500 (4%) |
| Median number of days from surgery to post-operative chest CT (median, IQR) | 5.5 [IQR 3-13] |
| Number of patients with confirmed COVID-19 from a post-operative viral swab | 4/41 (10%) |
| Median number of days from surgery to first symptom in those with confirmed COVID-19 | 5.5 [IQR 2-19] |
| Number of patients with chest CT showing typical changes of COVID-19<sup>a</sup> | 2/19 (11%) |
| Number of patients experiencing at least one clinical symptom that may be associated with COVID-19 | 47/500 (9%) |
| Cough | 21 |
| Fever | 29 |
| Shortness of breath | 25 |
| Muscle pain | 11 |
| Fatigue | 14 |
| Joint pain | 6 |
| Sore throat | 1 |
| Loss of smell | 3 |
| Loss of taste | 1 |
| Vomiting | 1 |
| Chest pain | 1 |
| Loss of appetite | 2 |
| Number of patients with probable COVID-19<sup>b</sup> |  |
| Number of patients with fever and at least one sign of acute respiratory illness | 6/500 (1%) |
| Median number of days from surgery to diagnosis of probable COVID-19 (median, IQR) | 14 ([QR 7-26]) |
| Number of patients with confirmed or probable COVID-19 | 10/500 (2%) |

<sup>a</sup>CT Chest with the typical appearances of COVID-19 pneumonia according to the Radiological Society of North America [19]
<sup>b</sup>A diagnosis of probable COVID-19 was given to patients who did not undergo laboratory testing or in whom laboratory testing was inconclusive, but who had fever and at least one sign of acute respiratory illness [18]

There were 92/500 (18%) grade 1-5 Clavien-Dindo complications, of which 33/500 (7%) were grade 3a or above (Table 5). The majority of these complications (32/33, 97%) were in patients without confirmed or probable COVID-19. One of these complications occurred in a patient with probable COVID-19. This was a grade 4b complication following an infected implant which required admission to ITU for management of septic shock and hypoxia. The patient was discharged home well on the 12^th^ post-operative day and developed probable COVID-19 on the 30^th^ post-operative day. They recovered fully at home without any treatment.

**Table 5:** Description of complications occurring within 30-days for Clavien-Dindo Grade 3 or above complications for 500 patients undergoing surgery:

| Clavien Dindo grade<sup>a</sup> | Complication | Frequency (n, %) |
| --- | --- | --- |
| IIIa<br>Requires surgical, endoscopic or radiological intervention under local anaesthetic | Anastomotic leak requiring urethral catheter<br>Urinary retention requiring catheterisation<br>Knee swelling requiring aspiration<br>Additional suture to improve seal of drain | n = 14 (3%)<br>1<br>11<br>1<br>1 |
| IIIb<br>Requires surgical, endoscopic or radiological intervention under general anaesthetic | Return to theatre due to post-operative bleeding | n = 2 (1%)<br>2 |
| IVa<br>Life-threatening complication requiring ITU management with single organ dysfunction | Admission to ITU for respiratory support following respiratory failure<br>Admission to ITU for cardiovascular support following post-operative bleed and/or hypotension<br>Admission to ITU for treatment of severe hyponatraemia<br>Admission to ITU for management of fast atrial fibrillation and haemodynamic compromise<br>Admission ITU for cardiac support following bradycardia and hypotension | n = 9 (2%)<br>3<br>3<br>1<br>1<br>1 |
| IVb<br>Life-threatening complication requiring ITU management with multi organ dysfunction | Admission to ITU for vasopressors for hypotension, intubated and ventilated for respiratory failure and treated for hyperkalaemia following acute kidney injury. | n = 5 (1%)<br>1 |
|  | Admission to ITU for cardiac support for right ventricular failure following cardiac arrest and respiratory support with non-invasive ventilation. | 1 |
|  | Admitted to ITU for intubation and ventilation after airway compromise from surgical emphysema and for vasopressors | 1 |
|  | Admission to ITU for respiratory support following hypoxia and supportive treatment for hepatic failure. | 1 |
|  | Admission to ITU for vasopressors for hypotension and high flow oxygen for hypoxia. | 1 |
| V<br>Death | Aspiration pneumonia | n = 3 (1%)<br>1 |
|  | Coronary atheroma due to underlying ischaemic heart disease | 1 |
|  | Metastatic breast cancer | 1 |

## Discussion

The principle finding of this study was that it is feasible and safe to continue with high-volume non-emergency surgery during the COVID-19 pandemic. No patient died from COVID-19 despite being in the peak of the pandemic in the worst affected region of the UK, which is a country with one of the highest number of cases and deaths from COVID-19 in the world [11, 23]. With an estimated 2 million surgeries being cancelled each week globally because of uncertainties associated with COVID-19 [2], patients are at risk of poorer survival outcomes and poorer quality of life [4-6]. This study has significant implications in supporting the continued provision of surgical services during the pandemic, gives a model for institutions wishing to continue performing surgery to follow and has implications for the surgical management of patients in future pandemics.

The 30-day mortality and complications from COVID-19 were much lower than those seen in previous studies, where mortality rates of 19-21% have been reported [7, 24], It is likely that these results reflected selection bias from only including patients with serious complications of COVID-19. Ten (2%) of the patients in the current cohort had probable or confirmed COVID-19 and none of these patients died from COVID-19. Overall a 7% Clavien-Dindo grade 3a or higher complication rate is a low rate of complications given the nature of surgeries being performed. This may reflect expertise at a high-volume tertiary cancer centre and patient selection. Patients were chosen who would benefit the most from surgery, balanced by their risk of serious complications from COVID-19. This is reflected in the overall patient demographics, which represent a relatively young, less co-morbid population than would typically have surgery at our institution. Importantly developing confirmed or probable COVID-19 infection did not appear to influence the likelihood of developing a complication.

Service reconfiguration was important in achieving the outcomes demonstrated. A hub-and-spoke model of practice was set up, with efforts on preserving the hub’s status as a COVID cold site. The hub accepted referrals from a multicentre surgical network, allowing the cases with the highest risk disease across different specialities within the network who would benefit most from surgery to be prioritised. Important local adjustments included diverting the majority of patient transfers or emergencies to an alternative geographically separate site within the institution. Footfall within the hospital was reduced by enabling staff to work from home when possible and for patient consultations to become telephone based.

PPE measures were introduced with the rationale of increasing the safety of staff and patients. Though some recommend universal operating room respiratory precautions in the pandemic [21] and this is what our institution adopted, there are uncertainties over this practice. For example, intubation and extubation during a general anaesthetic are aerosol-generating procedures that carry a higher risk of transmission of COVID-19, though there is less certainty over transmission risk from laparoscopy and from the production of a smoke plume from coagulating instruments. Performing surgery in full PPE is challenging, particularly during major and complex surgery, which comprised a large proportion of our cases. The impact on increasing the operative time and turnaround time between cases is not insignificant, meaning only a reduced surgical workload is feasible. Institutions should consider the implication that adopting these measures has on their ability to offer surgery during the peaks and recovery phases of the pandemic and further evidence to support the influence of these measures on risk of transmission of COVID-19 is warranted.

It is worth noting that measures such as pre-operative viral swabs and pre-operative CT chest testing were only introduced towards the end of this series, and despite this, the COVID-specific mortality rate remained low. This may suggest that other measures such as striving to maintain a COVID-free site, checking patients remained asymptomatic prior to their surgery and patient isolation pre and post-surgery could be the principle drivers of the observed outcomes.

There are a number of limitations in this study. First, not all of the patients were tested with a viral swab. This may underestimate the number of patients with confirmed laboratory diagnoses of COVID-19, though this may be mitigated by our assessment of patients for probable COVID-19 on the basis of their symptoms and in line with WHO guidelines [18]. Testing everyone in the community is not feasible in countries such as the UK, where testing capacity was limited, and government policy meant that testing was typically carried out for patients admitted to hospital.

Second, this service reconfiguration approach may not be feasible in all healthcare settings. At other institutions, particularly those based in one building, it may not be possible to keep the site COVID-free. However, we would strongly recommend that neighbouring institutions work together to designate cold COVID sites amongst a group of institutions during these unprecedented times.

Third, we should acknowledge the ethical dilemmas surrounding resource allocation at a time of limited resources and with uncertainty about where resources are best used [25]. The ability to offer such a service is dependent on local resources and the specific clinical situation, though models have been developed to allow planning for resource allocation during a pandemic [26]. It is ultimately down to the judgment of the regional healthcare system leaders whether it is appropriate and safe to offer the described approach.

## Conclusion

This study has demonstrated that it is feasible and safe to carry out non-emergency surgery during the COVID pandemic providing appropriate service reconfiguration takes place to facilitate this.

## Data Availability

The data that support the findings of this study are available upon reasonable request from bona fide researchers.

## Funding

Veeru Kasivisvanathan is an Academic Clinical Lecturer funded by the United Kingdom National Institute for Health Research (NIHR). The views expressed in this publication are those of the author(s) and not necessarily those of the NHS, the National Institute for Health Research or the Department of Health.

## Conflicts of Interests

The authors have no relevant conflicts of interest

## Notes

### Competing Interest Statement

The authors have declared no competing interest.

### Clinical Trial

This was not a clinical trial.

### Funding Statement

No direct study funding was received. However we would like to acknowledge: Veeru Kasivisvanathan is an Academic Clinical Lecturer funded by the United Kingdom National Institute for Health Research (NIHR). The views expressed in this publication are those of the author(s) and not necessarily those of the NHS, the National Institute for Health Research or the Department of Health.

### Author Declarations

The institutional review board at University College London Hospital deemed this work to be exempt from ethical approval.

